# Prognostic significance of the C-reactive protein–albumin–lymphocyte (CALLY) index in primary lung malignancies: a systematic review and meta-analysis with GRADE assessment

**DOI:** 10.64898/2026.10.18.26365819

**Authors:** Danyal Yarahmadi, Fatemeh Naseri Rad, Ihsaan Patel, Mohammad Yarahmadi, Aida Rashidi, Ali Yarahmadi, Parastou Kordestani Moghadam, Niayesh Naghavi, Mohammadsadra Shamohammadi

## Abstract

**Background:** The C-reactive protein–albumin–lymphocyte (CALLY) index integrates systemic inflammation, nutritional status, and immune function. We evaluated its association with survival in primary lung malignancies.

**Methods:** Embase, PubMed, Scopus, and Web of Science were searched from inception to 18 September 2026. Risk of bias was assessed using the Quality In Prognosis Studies tool. Hazard ratios (HRs) were synthesized using random-effects models with restricted maximum likelihood estimation and Hartung–Knapp adjustment. Sensitivity analyses, subgroup analyses, and meta-regression examined robustness and heterogeneity.

**Results:** Eleven studies contributed 13 cohorts to the overall survival (OS) analysis and 8 cohorts from seven studies to a combined recurrence/progression-free survival (RFS/PFS) analysis. Lower CALLY was associated with poorer OS (HR 1.78, 95% CI 1.49–2.12; I²=71.6%) and poorer RFS/PFS (HR 1.60, 95% CI 1.40–1.84; I²=0%). Associations persisted in leave-one-out analyses. No statistically significant subgroup differences were detected. Funnel-plot asymmetry suggested small-study effects for OS and RFS/PFS. Seven studies had moderate and four had high overall risk of bias. The certainty of evidence was rated as low for both OS and RFS/PFS, primarily due to risk of bias, heterogeneity, and endpoint indirectness.

**Conclusion:** Lower CALLY was associated with adverse survival outcomes in the current synthesis. Methodological limitations and variation in biomarker measurement and endpoint definitions restrict interpretation. Prospective validation using standardized measurements and clinically coherent outcomes is needed to establish incremental prognostic value and clinical utility.

## Introduction

Lung cancer (LC) is the most commonly diagnosed cancer worldwide and remains the leading cause of cancer-related mortality (1). According to GLOBOCAN 2024, approximately 2.6 million new cases and 1.9 million deaths are attributable to LC annually, with annual LC cases projected to reach approximately 4.6 million by 2050 (1, 2). Lung cancer is broadly classified into non-small cell lung cancer (NSCLC), accounting for approximately 85% of cases, and small cell lung cancer (SCLC), accounting for approximately 15%; these subtypes differ markedly in prognosis, clinical behavior, and treatment (3, 4). Given these differences, the Tumor-Node-Metastasis (TNM) staging system remains central to prognostic assessment and treatment planning; however, as an anatomical classification, it does not account for host-related factors such as systemic inflammation, nutritional status, and immunocompetence, which may contribute to differences in outcomes among patients with otherwise similar disease characteristics (5, 6).

Several blood-derived inflammatory and nutritional biomarkers have therefore been investigated as prognostic indicators in LC (7). These markers are inexpensive, routinely available, and easily calculated from standard laboratory parameters (7). Previous studies have evaluated the prognostic value of the neutrophil-to-lymphocyte ratio (NLR), platelet-to-lymphocyte ratio (PLR), Prognostic Nutritional Index (PNI), C-reactive protein-to-albumin ratio (CAR), and Glasgow Prognostic Score or modified Glasgow Prognostic Score (mGPS) (8–11). These indices capture different aspects of the host response to malignancy and have demonstrated prognostic value across a range of LC populations and treatment settings (7–11).

The C-reactive protein–albumin–lymphocyte (CALLY) index is a multicomponent biomarker incorporating serum C-reactive protein as a marker of inflammation, albumin as a marker of nutritional status, and peripheral lymphocyte count as a marker of immune status (12). Initially developed as a prognostic biomarker for hepatocellular carcinoma, CALLY has subsequently been evaluated in LC (12, 13). In a cohort of 1,864 patients with NSCLC, low CALLY was associated with poorer overall survival (OS), while a CALLY-based prognostic model demonstrated greater prognostic discrimination than TNM staging alone (13). Subsequent studies in surgically treated NSCLC populations similarly reported associations between lower CALLY values and poorer OS and recurrence-related outcomes, with findings also reproduced in discovery and external validation cohorts (14–16). Beyond surgical populations, post-treatment CALLY has demonstrated prognostic value in patients with locally advanced NSCLC receiving thoracic radiotherapy (17).

Despite these findings, the prognostic role of CALLY in LC remains incompletely defined (18). Existing studies differ in disease stage, patient population, treatment setting, timing of CALLY assessment, cutoff determination, and survival outcomes, limiting direct comparison and generalizability (13–18). Much of the available evidence is retrospective and predominantly relates to NSCLC, with several studies focusing specifically on surgically treated populations (13–18). Accordingly, this systematic review and meta-analysis evaluated the prognostic significance of the CALLY index and its association with survival outcomes in patients with LC.

## Methods

This systematic review and meta-analysis evaluated the prognostic significance of the CALLY index in primary lung malignancies and was reported in accordance with PRISMA 2020 (19). The review protocol was registered in PROSPERO under registration number CRD420261475710.

### Information sources and search strategy

Embase, PubMed, Scopus, and Web of Science were searched from database inception to 18 September 2026 to identify studies evaluating the prognostic significance of the CALLY index in primary lung malignancies. The search combined terms for the CALLY index, including “CALLY,” “CALLY index,” and “C-reactive protein–albumin–lymphocyte,” with lung cancer terms such as “lung cancer,” “lung neoplasm,” “non-small cell lung cancer,” “NSCLC,” “small cell lung cancer,” and “SCLC.” Search syntax was adapted to each database using controlled vocabulary where available and free-text terms. Complete database-specific strategies, search dates, and applied restrictions are provided in Supplementary Table S2.

### Eligibility criteria

Studies were eligible if they evaluated the CALLY index in adults with a primary lung malignancy and reported its association with a time-to-event survival outcome. Non-small cell and small cell lung cancers were considered eligible. Studies contributing to meta-analysis were required to report an HR with a 95% CI or sufficient information to derive these quantities. Reports concerning secondary pulmonary malignancies, nonclinical studies, case reports, and publications without extractable prognostic data were excluded. Multiple reports from the same population were evaluated together to avoid counting overlapping participants more than once. Distinct discovery and validation cohorts were considered separate analytical units when their independence could be established.

### Data collection process and data items

Data extraction covered publication details, study setting and design, recruitment period, cohort size, demographic characteristics, histology, disease stage, treatment, and follow-up. Biomarker data included the CALLY formula and laboratory units, sampling time, modeling scale, cutoff, and method of cutoff selection. For each outcome, the definition, time origin, event count where available, HR, 95% CI, comparison direction, adjustment level, and covariates were recorded. The exact source table or figure was retained to permit verification. Discovery and validation cohorts were distinguished, and potential population overlap was examined. Given the limited number of studies reporting recurrence-free survival (RFS) and progression-free survival (PFS) separately, these outcomes were combined to preserve statistical precision and avoid unstable estimates. Although their definitions are not identical, both capture recurrence- or progression-related disease control. Similar harmonization of related survival endpoints has been adopted in previous oncology meta-analyses (20, 21). Nevertheless, differences in endpoint definitions and clinical settings across the included studies were acknowledged as potential sources of heterogeneity, warranting cautious interpretation.

For studies without directly reported categorical HRs (17, 22), unadjusted HRs and corresponding 95% confidence intervals were reconstructed from digitized Kaplan–Meier survival curves following the methodological framework described by Tierney et al. (2007), incorporating reported numbers at risk where available and assumptions regarding censoring (23).

### Risk of bias assessment

Risk of bias was assessed using QUIPS (24) across study participation, study attrition, prognostic factor measurement, outcome measurement, study confounding, and statistical analysis and reporting. Each domain was judged as low, moderate, or high risk. The assessment considered cohort selection, completeness of follow-up, the timing and consistency of CALLY measurement, outcome ascertainment, adjustment for established prognostic factors, and the appropriateness of statistical modeling.

### Certainty of evidence assessment

Certainty was assessed separately for each clinically coherent outcome using GRADE guidance for prognostic factors (25). Under this framework, observational prognostic evidence starts at high certainty and is assessed for risk of bias, inconsistency, indirectness, imprecision, and publication bias. QUIPS findings should inform risk-of-bias judgments. Inconsistency should consider clinical comparability and the distribution of effects, while indirectness should consider applicability to the target population, biomarker timing, and outcome. Imprecision should consider confidence intervals and information size. Small-study effects were assessed alongside heterogeneity and the number of available studies.

### Statistical analysis

Hazard ratios (HRs) with 95% confidence intervals (CIs) were used as the summary effect measure for overall survival (OS) and RFS/PFS. Reported HRs were log-transformed, and standard errors were reconstructed from the corresponding 95% CIs. Pooled estimates were generated using generic inverse-variance random-effects models, with between-study variance estimated by restricted maximum likelihood (REML) and Hartung–Knapp adjustment applied to random-effects inference. Statistical heterogeneity was quantified using Cochran’s Q, I², and τ².

Small-study effects were assessed using contour-enhanced funnel plots centered on the null effect, together with Egger’s regression and Begg–Mazumdar rank-correlation tests; these tests were interpreted cautiously when fewer than 10 studies were available. Robustness of pooled estimates was examined through leave-one-out analyses, refitting the REML–Hartung–Knapp model after sequential omission of each study.

For OS, prespecified subgroup analyses examined sample size (<500 vs >500), CALLY cutoff (<2 vs >2), and adjustment level (multivariable vs univariable), assuming a common residual τ² across subgroup levels and using formal tests for subgroup differences. Univariate meta-regression further evaluated sample size, adjustment level, CALLY cutoff, and male and female participant counts as study-level moderators. Sample size and sex counts were scaled per 100 participants, whereas cutoff was modeled per unit increase. Moderator-specific complete-case analyses were used where data were missing. Analyses were conducted in R version 4.6.1 using the *meta* and *metafor* packages.

## Results

### Study selection process

The searches identified 1,046 records: 247 from Embase, 122 from PubMed, 473 from Scopus, and 204 from Web of Science. After removal of 313 duplicates, 733 records underwent title and abstract screening, and 34 reports were assessed in full text. Eleven studies describing 13 cohorts met the review eligibility criteria. The selection flow and reasons for full-text exclusion are presented in Figure 1.

**Figure 1.**
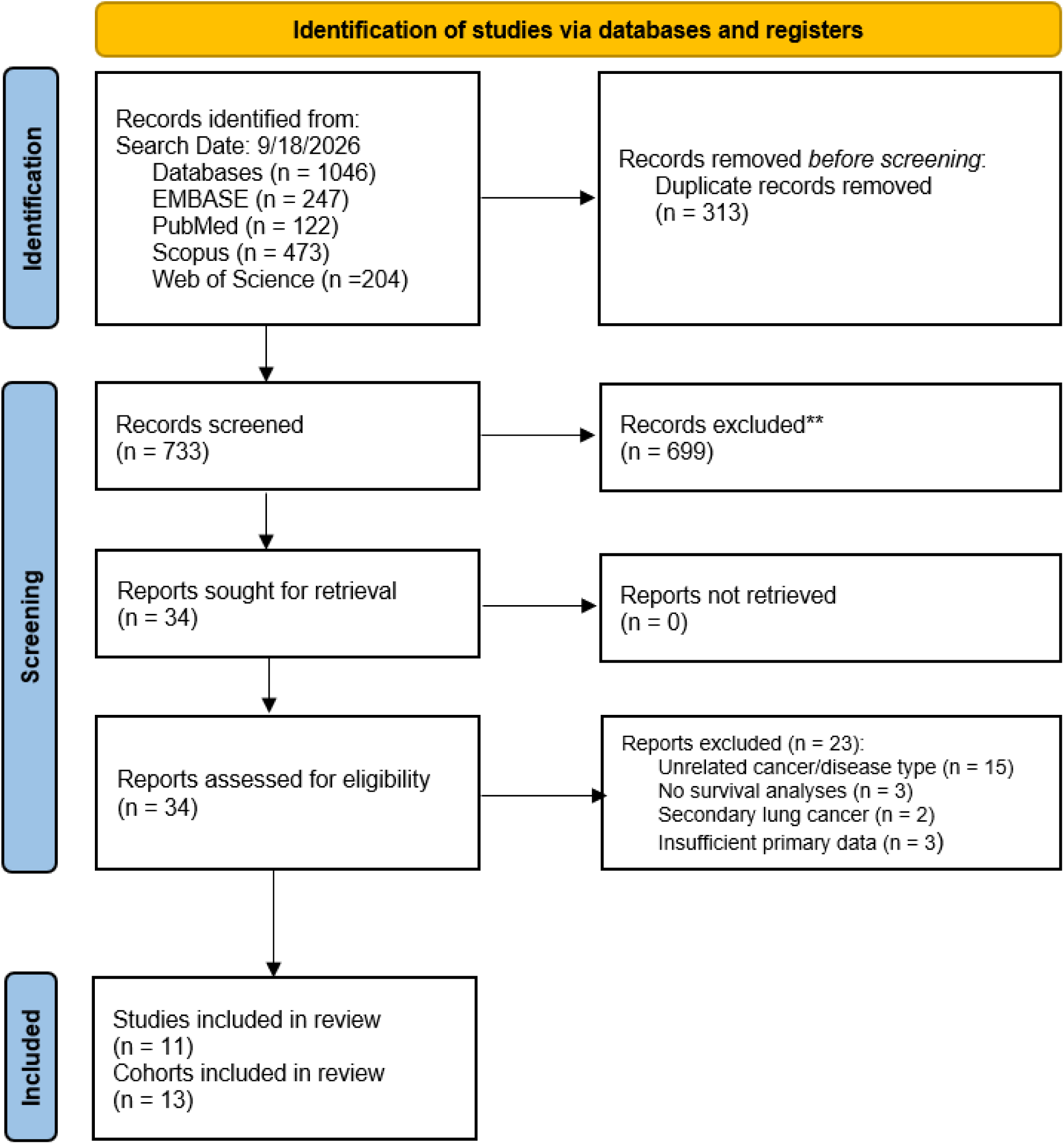
PRISMA 2020 flow diagram of the study selection process.

### Baseline characteristics

Eleven studies published between 2023 and 2026 described 13 cohorts comprising 5,283 participants (13–17, 22, 26–30). Three studies were conducted in China, three in Japan, four in Turkey, and one in South Korea. Individual cohort sizes ranged from 81 to 1,307 participants. Approximately 65% of participants were male. Five studies evaluated patients undergoing lung cancer resection, including a cohort restricted to patients with interstitial pneumonia. The remaining studies examined mixed-stage NSCLC receiving various treatments, unresectable stage III NSCLC treated with thoracic radiotherapy, advanced NSCLC receiving first-line immunotherapy-based treatment, metastatic NSCLC treated with nivolumab in the second-line or later setting, extensive-stage SCLC treated with platinum–etoposide chemotherapy, and older adults undergoing radiotherapy. Thresholds were commonly selected using receiver operating characteristic analyses, maximally selected statistics, or cohort medians. Study characteristics are summarized in Table 1.

**Table 1.** Characteristics of included studies.

| Study and cohort | Country design and recruitment | N | Age years | Female / male | Population and treatment | Cutoff and selection method | Low / high N | Follow-up | Reported outcome |
| --- | --- | --- | --- | --- | --- | --- | --- | --- | --- |
| Liu 2023 Training (13) | China; multicentre<br>Retrospective analysis of prospective registry<br>May 2013–Dec 2018 | 1307 | 61 (54–67) | 454/853 | NSCLC; I–IV<br>Mixed treatment; surgery 289, chemotherapy 804, RT 70 | 1.32; low ≤1.32<br>Maximally selected rank statistics | 748/559 | 21.31 (17.67)‡ | OS |
| Liu 2023 Validation (13) | Same registry; random split<br>May 2013–Dec 2018 | 557 | 61 (54–67) | 199/358 | NSCLC; I–IV<br>Surgery 122, chemotherapy 334, RT 24 | 1.32; same threshold | 316/241 | 22.33 (19.47)‡ | OS |
| Cheng 2025 (14) | China; single centre<br>Retrospective<br>Feb 2008–Mar 2013 | 302 | ≥60: 152 (50.3%) | 95/207 | NSCLC; pStage I–III<br>R0 resection; no prior treatment | 1.38; low ≤1.38<br>X-tile for OS | 117/185 | Minimum 3 years; final Mar 2016 | OS |
| Şeber 2025 (29) | Turkey; single centre<br>Retrospective<br>2012–2024 | 167 | 62 (40–88)† | 14/153 | Extensive-stage SCLC<br>First-line platinum–etoposide; no immunotherapy | 4.55; low $<$4.55<br>Median, after unsuccessful ROC selection | 112/55 | NR | OS |
| Mizota 2026 (15)<br>Discovery KUH | Japan; retrospective<br>2013–2018 | 581 | 70 (65–76) | 241/340 | NSCLC; cStage I–IIIA<br>Resection; adjuvant chemotherapy 61 | 6.2; low $<$6.2<br>Time-dependent ROC for 5-year OS | 252/329 | Median NR; 5-year analyses | RFS / OS |
| Mizota 2026<br>(15)<br>Validation<br>KCC | Japan; external centre<br>Retrospective<br>2013–2018 | 634 | 69 (63–<br>76) | 287/347 | NSCLC; cStage I–IIIA<br>Resection; adjuvant<br>chemotherapy 199 | 6.2; same<br>threshold | 281/353 | Median NR; 5-year analyses | RFS / OS |
| Kim 2026<br>(27) | South Korea; single<br>centre<br>Retrospective<br>Jan 2011–Dec 2020 | 680 | 66 (59–<br>73) | 276/404 | NSCLC; pStage Tis/I–III<br>Curative-intent resection;<br>neoadjuvant therapy excluded | 5.14; low ≤5.14<br>Maximally<br>selected log-rank<br>statistics for 5-year OS | 273/407 | Median NR; 5-year analyses | RFS / OS |
| Konoeda<br>2026 (28) | Japan; single centre<br>Retrospective<br>Jan 2011–Mar 2021 | 85 | 73 (66–<br>78) | 20/65 | Lung cancer with interstitial<br>pneumonia; cStage I–III; pStage<br>0–III<br>Resection | 0.97; low ≤0.97<br>ROC for<br>postoperative OS | 20/65 | Median 1112<br>days | RFS / OS |
| Yoneyama<br>2026 (16) | Japan; single centre<br>Retrospective<br>May 2016–Aug 2023 | 329 | 72 (67–<br>76) | 119/210 | Invasive NSCLC; pStage I–III<br>Curative resection; neoadjuvant<br>therapy excluded | 6.53; low <6.53<br>ROC/Youden for<br>2-year RFS | 166/163 | Median 54 (35–<br>69) months | RFS / OS |
| Akkar 2026<br>(22) | Turkey; single centre<br>Retrospective<br>Aug 2020–Dec 2024 | 81* | NA | NA | NSCLC ; SCLC<br>RT; palliative intent | 0.50; low <0.50<br>ROC for mortality | 57/24 | Median 8 (0–43)<br>months after RT | OS |
| Yin 2026<br>(17) | China; single centre<br>Retrospective<br>Jan 2012–Nov 2018 | 218<br>* | 61 (56–<br>65) | 43/175 | Unresectable NSCLC; IIIA–IIIC<br>Thoracic RT; concurrent<br>chemotherapy 88 | 0.63; median for<br>KM only<br>No baseline cutoff | 108/110§ | Median 46.3<br>(42.9–51.2)<br>months | Local PFS / OS |
| Bedir 2026<br>(26) | Turkey; three centres<br>Retrospective<br>Jan 2019–Jan 2025 | 161 | 63 (34–<br>82)† | 32/129 | Advanced NSCLC; de novo<br>metastatic 129, recurrent 32<br>First-line ICI alone 74, chemo-ICI<br>65, dual ICI 22 | 0.0275<br>ROC/Youden for<br>vital status at last<br>follow-up | 71/90 | Median 16.98<br>(1.84–112.85)<br>months† | OS / PFS |
| Günaltılı<br>2026 (30) | Turkey; two centers<br>Retrospective<br>May 2016–Oct 2024 | 181 | 64 (59-70) | 40/141 | Metastatic NSCLC receiving<br>Nivolumab monotherapy as<br>second or third line of therapy | 0.32<br>median value | 91/90 | Median 27.6<br>(22.3–32.6)<br>months | OS / PFS |

Age is median (IQR) unless marked †, indicating median (range). Cheng reports age categories. NR, not reported; UV, univariable; MV, multivariable; KM, Kaplan–Meier; PSM, propensity score matching; ICI, immune checkpoint inhibitor; RT, radiotherapy; NSCLC, non-small cell lung cancer; SCLC, small cell lung cancer; OS, overall survival; RFS, recurrence-free survival; PFS, progression-free survival; LPFS, locoregional progression-free survival; DMFS, distant metastasis-free survival; NA, not available. ‡ Liu reports mean follow-up (SD), in months. † Range. Other three-number follow-up summaries reproduce the source without inferring an unspecified interval type. § Yin group counts refer to post-treatment CALLY Kim counts refer to the original 680 patients; PSM yielded 208 patients per group. \* HRs and 95% CIs were reconstructed from Kaplan–Meier curves using the methods described by Tierney et al..

### Risk of bias assessment

The QUIPS assessment showed a heterogeneous methodological profile across the 11 included studies (Figure 6). No study was judged to have an overall low risk of bias; seven were classified as moderate risk and four as high risk. Study participation was rated as low risk in five studies, moderate in three, and high in three. Attrition was a more frequent concern, with only two studies rated low risk, six moderate, and three high. Prognostic factor measurement was consistently robust: ten studies were judged low risk and one moderate, reflecting the largely objective laboratory-based derivation of the CALLY index. Outcome measurement was also a strong domain, with only one study rated as moderate risk. Confounding represented the principal methodological concern. Only one study was rated low risk in this domain, seven were moderate, and three were high, largely because of incomplete adjustment for clinically relevant prognostic factors or the potential for residual confounding. Statistical analysis and reporting were also commonly rated as moderate risk, with ten studies in this category and only one judged low risk. Overall high risk of bias was assigned to Cheng et al. (14), Bedir et al.(26), Şeber et al. (29), and Yin et al. (17); the remaining seven studies were judged to have moderate overall risk.

### Survival endpoints meta-analyses

Thirteen cohorts from 11 studies (13–17, 22, 26–30) contributed to the OS meta-analysis. Lower CALLY was associated with poorer OS (HR 1.78, 95% CI 1.49–2.12), with substantial heterogeneity (I²=71.6%; τ²=0.0581; Figure 2A). Eight cohorts from seven studies (15–17, 26–28, 30) contributed to the RFS/PFS synthesis (HR 1.60, 95% CI 1.40–1.84; I²=0%; Figure 2B).

**Figure 2.**
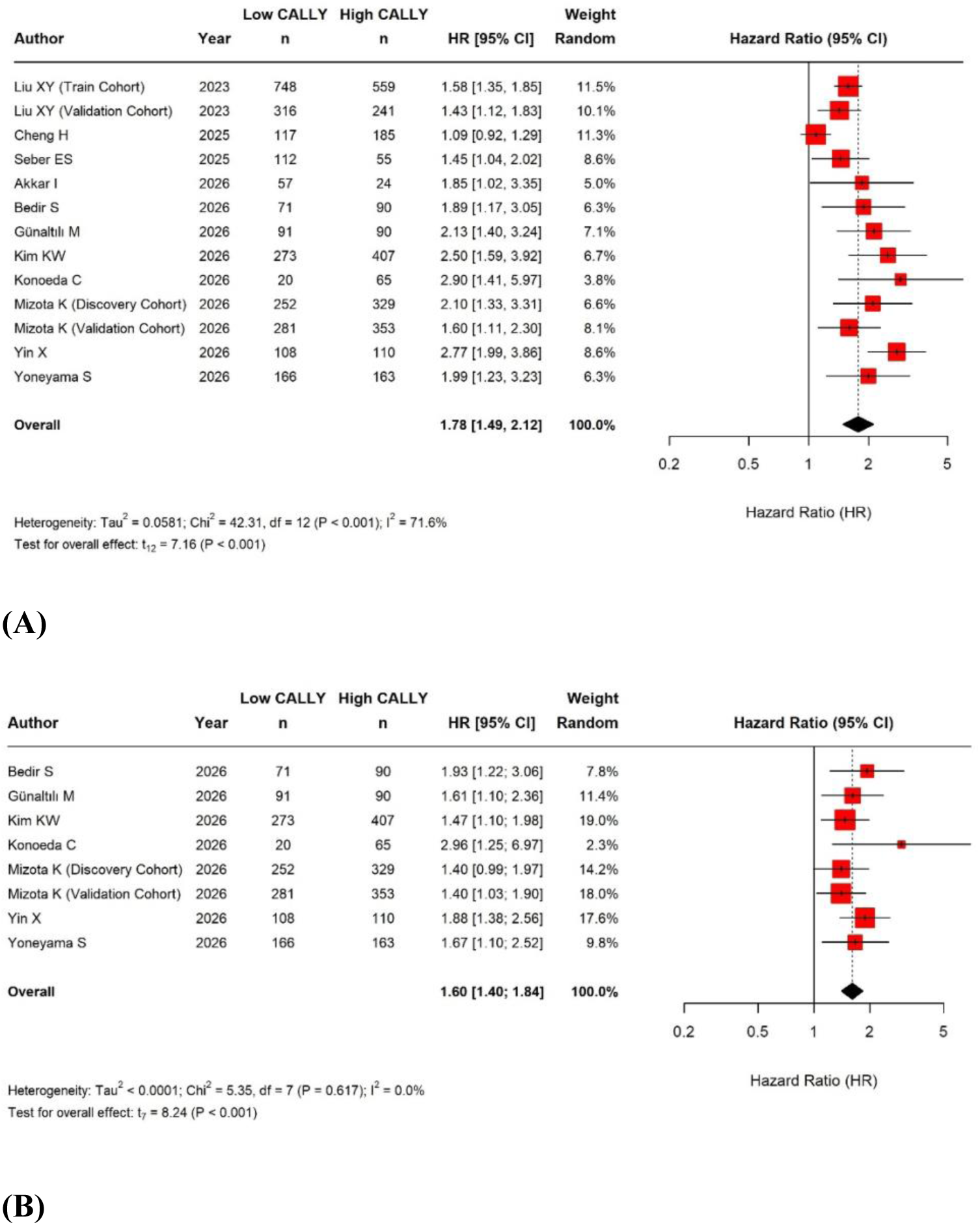
Forest plot of the association between the CALLY index and survival endpoints in lung cancer. (A) Overall survival and (B) RFS/PFS.

### Small-study effects and publication bias

For OS (11 studies; 13 cohorts), Egger’s test suggested funnel-plot asymmetry (P=0.011), whereas Begg’s test was not statistically significant (P=0.113; Figure 3A). Substantial heterogeneity limits attribution of this pattern to publication bias alone. For the combined RFS/PFS outcome, Egger’s test approached statistical significance (P=0.052), while Begg’s test indicated marginal asymmetry (P=0.048; Figure 3B). With only eight cohorts, these results were considered exploratory and insufficient to establish publication bias. Overall, these discordant findings suggest possible small-study effects but provide insufficient evidence to establish publication bias.

**Figure 3.**
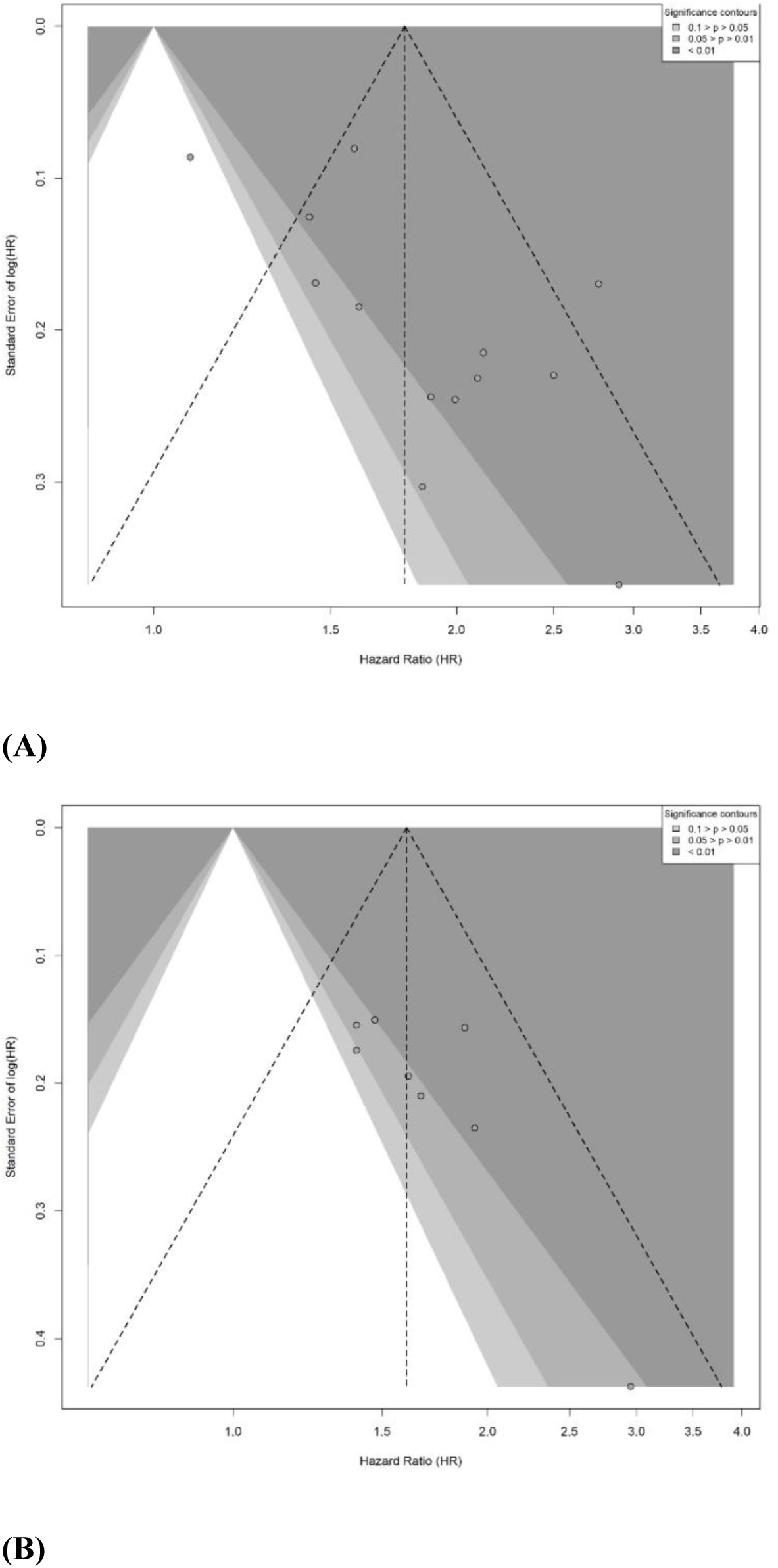
Contour-enhanced funnel plot for survival endpoints in the primary CALLY–lung cancer meta-analysis. (A) Overall survival and (B) RFS/PFS: Egger’s p = 0.011, Begg’s p = 0.113; (B) RFS/PFS: Egger’s p = 0.052, Begg’s p = 0.048.

### Sensitivity analyses

Leave-one-out analyses confirmed the stability of the pooled estimates for both survival outcomes. For OS, sequential omission of individual cohorts produced pooled HRs ranging from 1.68 to 1.86, and all corresponding 95% CIs remained above the null (Figure 4A). The largest absolute change from the full-model estimate occurred after exclusion of Yin X et al. (17), resulting in a 5.3% decrease in the pooled HR. No single cohort materially altered the direction or statistical significance of the association, indicating that the OS finding was not driven by an isolated influential study.

**Figure 4.**
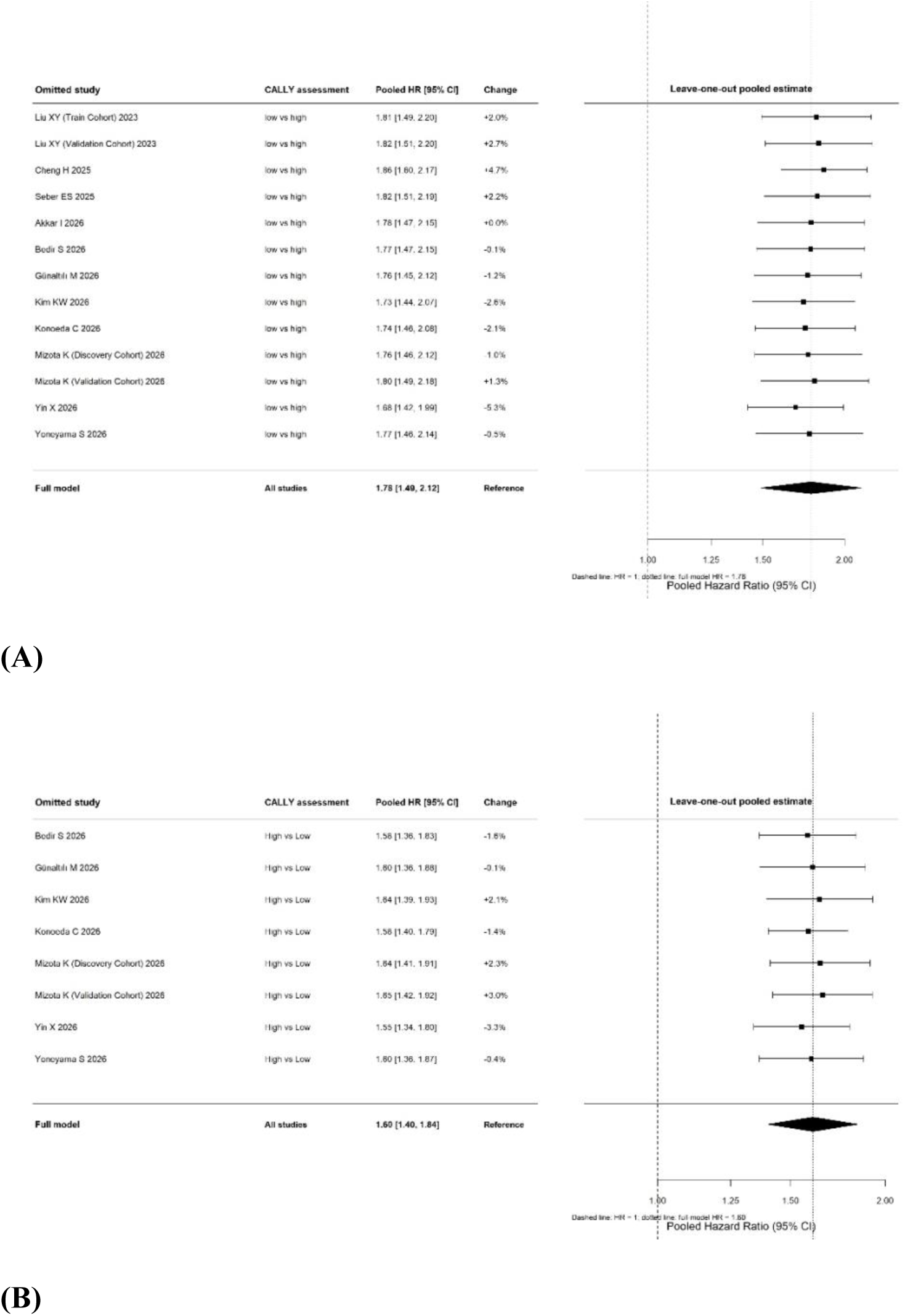
Leave-one-out influence analysis of the association between the CALLY index and survival endpoints in lung cancer. (A) Overall survival; (B) RFS/PFS.

The RFS/PFS analysis showed a similarly robust pattern (Figure 4B). Leave-one-out pooled HRs ranged from 1.55 to 1.65, with all 95% CIs excluding 1. The greatest absolute change was 3.3% following omission of Yin et al. (17). Across all iterations, the association between lower CALLY and poorer survival remained statistically significant. These findings support the robustness of the primary OS and RFS/PFS estimates to exclusion of individual cohorts.

### Subgroup analyses

The association between lower CALLY and poorer OS was numerically stronger in cohorts using cutoffs >2 (HR 1.86, 95% CI 1.41–2.44) than in those using cutoffs ≤2 (HR 1.74, 95% CI 1.32–2.30), although the subgroup difference was not statistically significant (P=0.678; Figure 5A). No statistically significant subgroup differences were detected by cohort size (P=0.793; Figure 5B) or adjustment level (P=0.504; Figure 5C). Within-subgroup heterogeneity was descriptively lower among cohorts using cutoffs >2 and those reporting multivariable-adjusted estimates.

**Figure 5.**
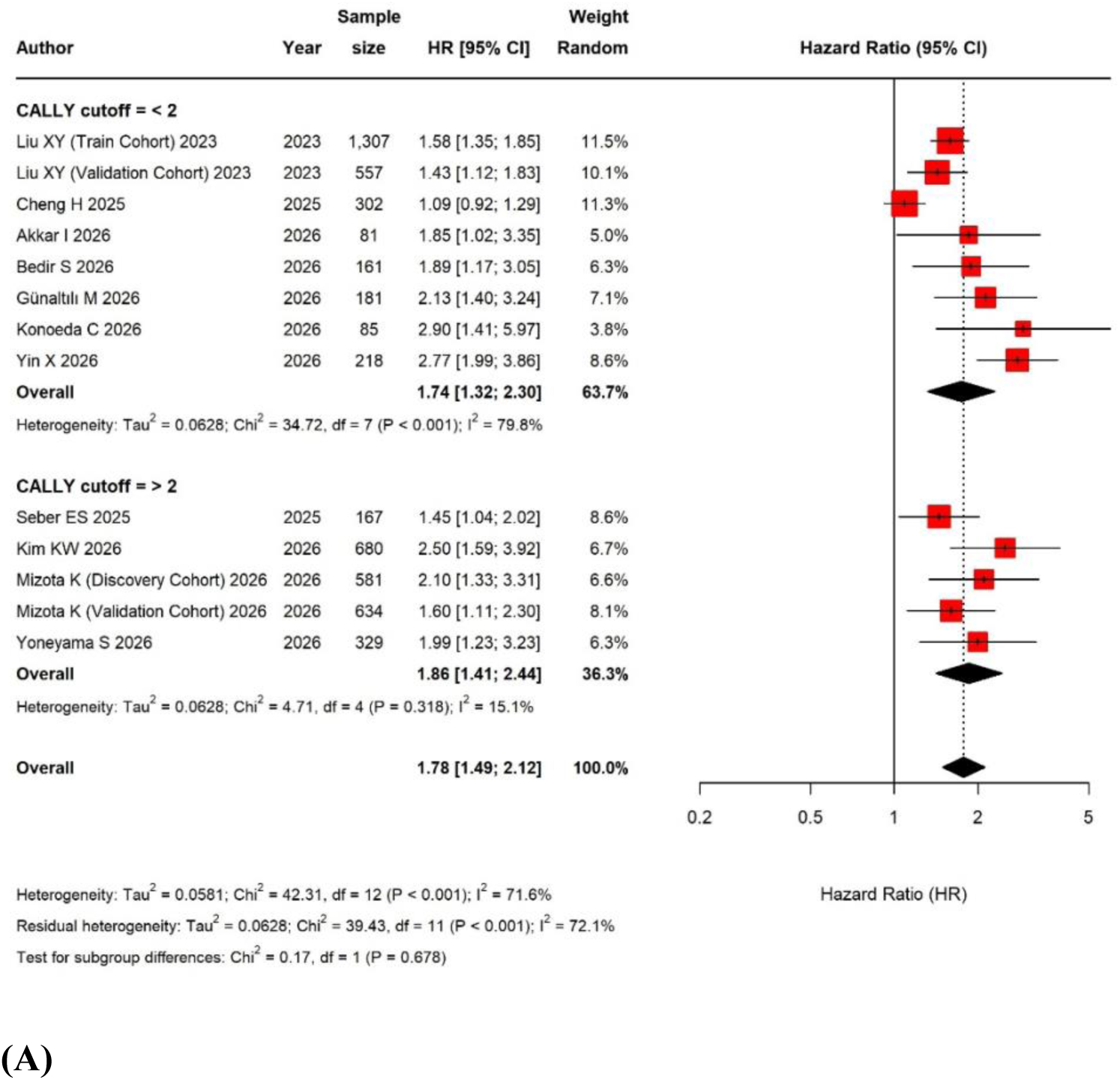
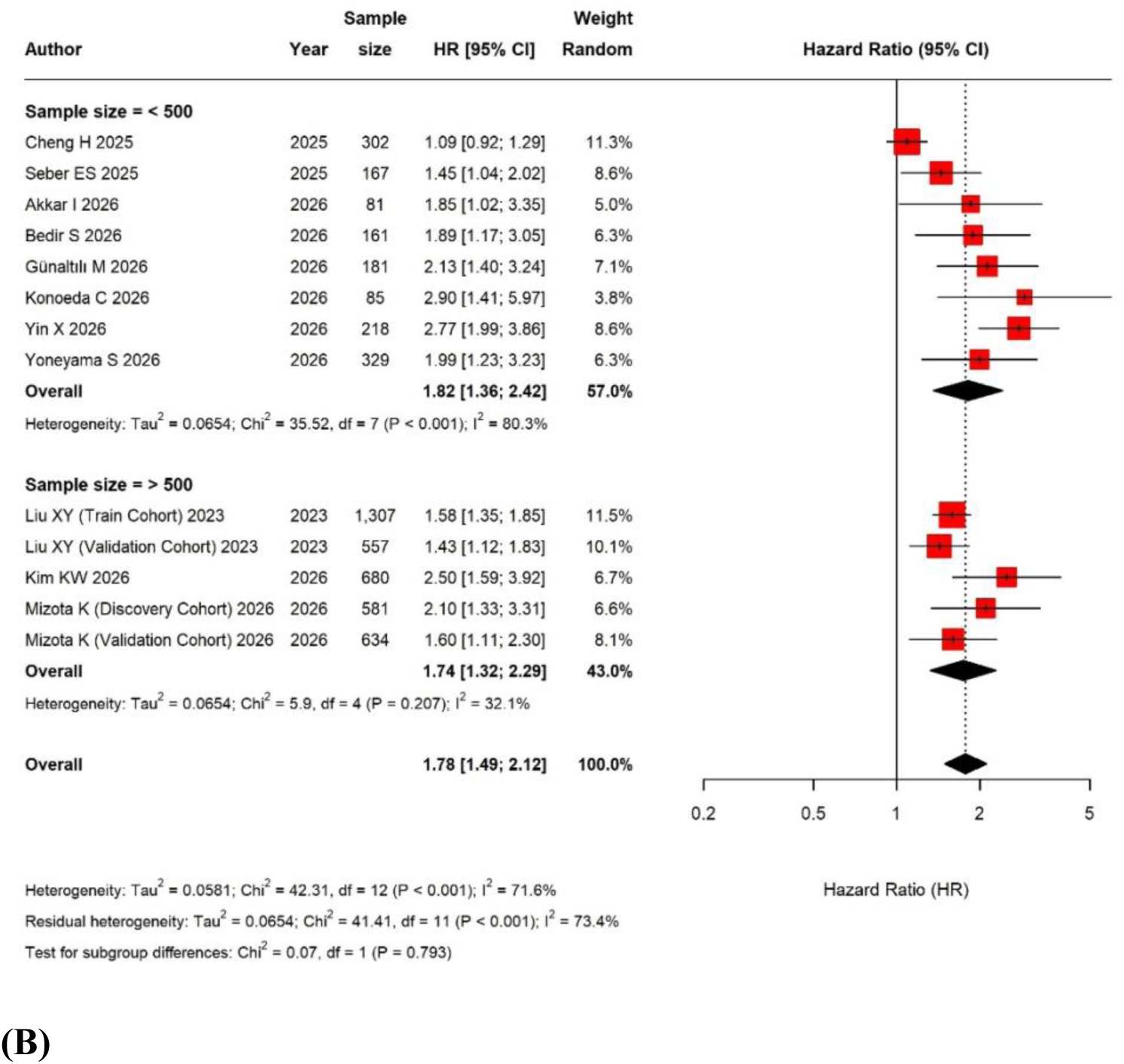
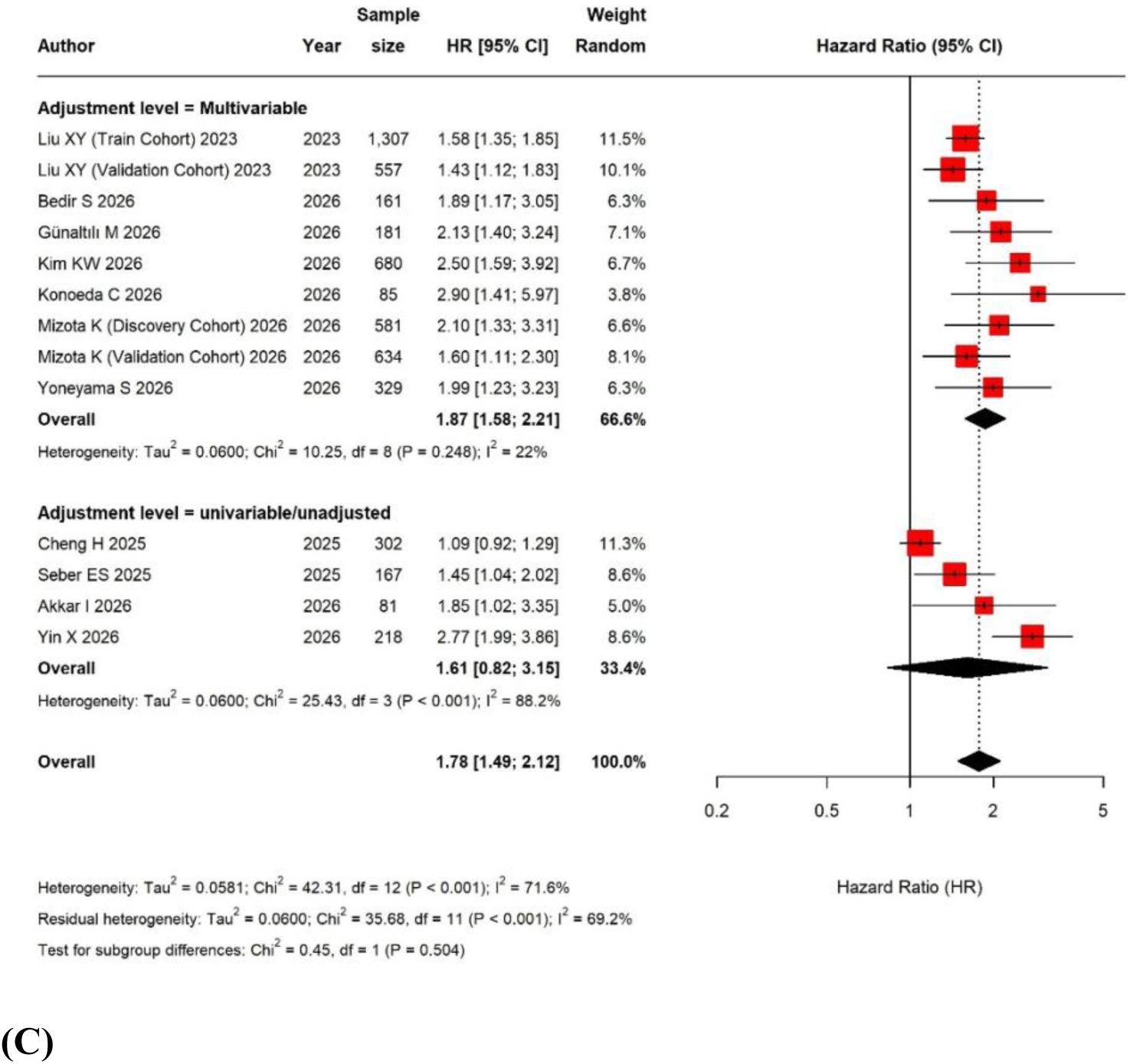
Subgroup analysis of the association between the CALLY index and overall survival. (A) according to the CALLY cutoff; (B) according to the sample size; (C) according to the adjustment level.

**Figure 6.**
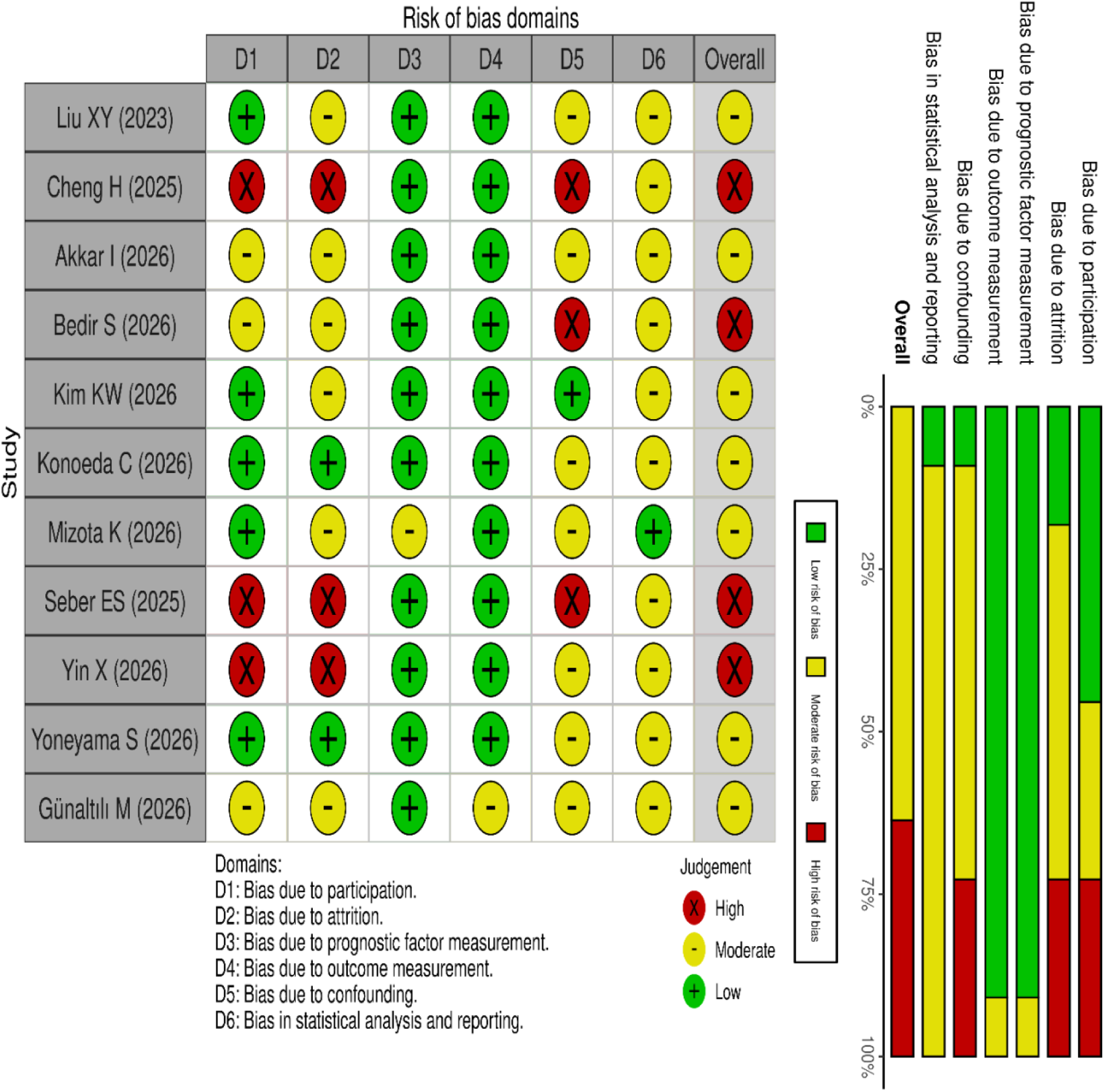
Risk-of-bias assessment of the included studies using the Quality In Prognosis Studies (QUIPS) tool. The upper panel presents study-specific judgments across six domains and the overall risk-of-bias judgment. The lower panel summarizes the percentage of studies assigned to each risk category. Domains comprise study participation (D1), study attrition (D2), prognostic factor measurement (D3), outcome measurement (D4), study confounding (D5), and statistical analysis and reporting (D6). Green (+), yellow (−), and red (×) indicate low, moderate, and high risk of bias, respectively.

### Meta-regression

In the univariable meta-regression analyses, none of the evaluated moderators was statistically significant (Table 2). The cutoff coefficient corresponded to a multiplicative change in HR of 1.004 per unit increase in the cutoff (P=0.906), with residual I² of 71.8%. Substantial residual heterogeneity persisted, and these exploratory study-level analyses did not establish a source of effect modification.

**Table 2.** Univariate meta-regression results for overall survival.

| Moderator | Unit | k | Beta | SE | Exp Beta | Moderator P | Parent I<sup>2</sup> | Residual I<sup>2</sup> |
| --- | --- | --- | --- | --- | --- | --- | --- | --- |
| Sample size | Per 100 additional participants | 13 | -<br>0.0136 | 0.0230 | 0.987 | 0.567 | 71.6% | 67.5% |
| Adjustment level | Univariable versus Multivariable | 13 | -<br>0.1486 | 0.1719 | 0.862 | 0.406 | 71.6% | 67.4% |
| CALLY cutoff value | Per 1-unit increase in cutoff | 13 | 0.0042 | 0.0352 | 1.004 | 0.906 | 71.6% | 71.8% |
| Male participant count | Per 100 additional male participants | 12 | -<br>0.0246 | 0.0403 | 0.976 | 0.556 | 73.8% | 70.2% |
| Female participant count | Per 100 additional female participants | 12 | -<br>0.0301 | 0.0632 | 0.970 | 0.644 | 73.8% | 70.8% |

### Certainty of evidence assessment

The certainty of evidence was rated as low for both OS and the combined RFS/PFS outcome (Table 3). Both outcomes were downgraded for serious risk of bias. OS was further downgraded for serious inconsistency due to substantial unexplained heterogeneity (I²=71.6%), whereas RFS/PFS was downgraded for serious indirectness arising from the combination of clinically distinct survival endpoints. No downgrades were applied for imprecision or publication bias, given the relatively precise pooled estimates and inconclusive evidence of small-study effects.

**Table 3.**
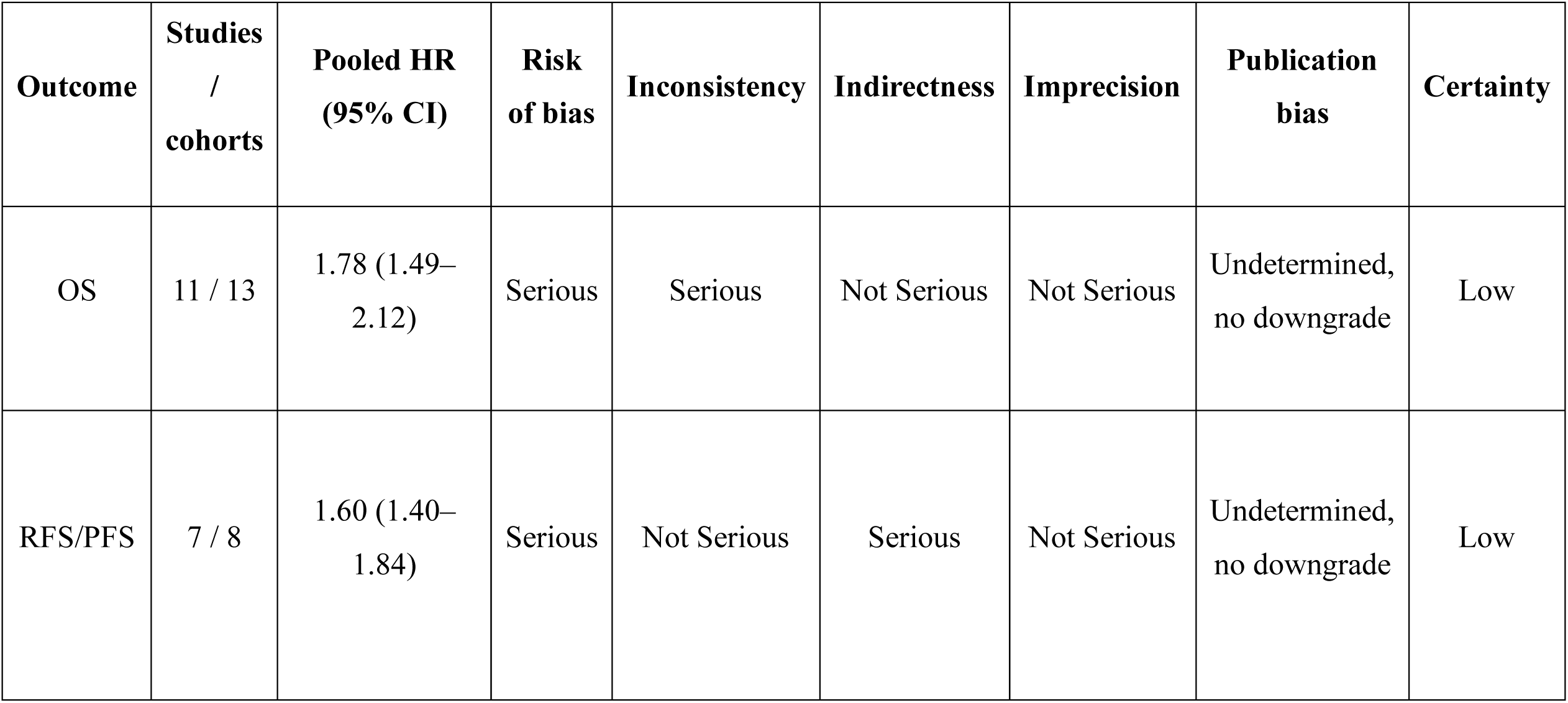
GRADE certainty of evidence assessment.

## Discussion

The current synthesis included 13 cohorts from 11 studies and found an association between lower CALLY and poorer OS. The pooled HR was 1.78, but substantial heterogeneity indicates that the magnitude of the association varied across populations and study methods. The RFS/PFS analysis also suggested an adverse association, with little observed statistical heterogeneity; nevertheless, differences in endpoint definitions limit its outcome specificity. Leave-one-out analyses did not identify a single cohort responsible for either pooled finding. Subgroup analyses did not demonstrate statistically significant differences, and meta-regression did not identify a statistically significant study-level moderator. These findings support further investigation of CALLY as a prognostic factor, while methodological limitations preclude treating the pooled associations as evidence of established clinical utility.

The present findings are broadly consistent with an expanding body of evidence positioning CALLY as a host-response marker rather than a tumor-specific biomarker. In surgically resected NSCLC, Mizota et al. showed in independent discovery and validation cohorts that low CALLY predicted both poorer RFS and OS (15). Kim et al. likewise found CALLY independently associated with OS and superior to the other evaluated systemic markers for OS discrimination, although RFS did not differ significantly between CALLY groups (27). This endpoint-specific inconsistency is informative. In a higher-risk surgical population with interstitial pneumonia, Konoeda et al. found low CALLY independently associated with poorer RFS and OS (28), while Yoneyama et al. demonstrated that low CALLY and sarcopenia were independent, complementary adverse prognostic factors after NSCLC resection (16). Yin et al. extended this signal beyond surgery: post-treatment CALLY independently predicted OS, LPFS, and DMFS in LA-NSCLC treated with thoracic RT (17).

Broader evidence points in the same direction. A pan-cancer meta-analysis and meta-analyses of colorectal and hepatobiliary and pancreatic cancers associated higher pretreatment CALLY with more favorable survival outcomes across multiple time-to-event endpoints (18, 31, 32). Parallel lung-cancer studies of other blood-based host-response indices reinforce this biological framework: LIPI stratified ICI outcomes, MLR independently predicted recurrence in stage IA NSCLC, and NLR strongly discriminated NSCLC from healthy controls (33–35). Our meta-analysis strengthens this disease-specific evidence, showing that low CALLY was associated with poorer OS and RFS, while cutoff-related analyses suggest that threshold selection may contribute to OS heterogeneity.

The principal clinical appeal of CALLY is its ability to summarize three clinically relevant host domains, systemic inflammation, nutritional reserve, and immune competence, from routine laboratory measurements. In resected NSCLC, Kim et al. reported that CALLY had the highest time-dependent AUC for OS among the compared inflammation-, nutrition-, and immunity-based markers (27). Evidence from other cancers also suggests that CALLY can add information beyond conventional staging. In nonmetastatic CRC, Lee et al. found that only CALLY and CAR remained independent among seven immunonutritional markers, and adding CALLY to stage increased the C-index from 0.713 to 0.812 (36). Complementarity may be equally important: Yoneyama et al. observed essentially no correlation between CALLY and skeletal muscle index, yet both independently predicted outcomes, supporting multidimensional rather than single-marker risk assessment (16).

CALLY may also reflect clinically relevant vulnerability beyond long-term survival. Low CALLY was associated with postoperative pneumonia and longer hospitalization in lung cancer with interstitial pneumonia (28), with inferior treatment response and greater relapse risk in DLBCL (37), and with poorer PFS and postoperative lung metastasis in CRC (38). Nevertheless, biomarker performance is context-dependent; Mezquita et al. showed that LIPI stratified outcomes with ICI but not chemotherapy (33), illustrating why host-response indices should not be interpreted independently of treatment context. Contemporary review evidence likewise emphasizes unresolved standardization and implementation challenges (39). CALLY warrants evaluation alongside established prognostic factors, including disease stage, performance status, comorbidity, and treatment context. Although the observed associations support further investigation, this meta-analysis does not establish incremental predictive value or benefit from CALLY-guided management. Substantial OS heterogeneity and possible small-study effects further support cautious interpretation.

The evidence was predominantly observational, and incomplete control of established prognostic factors leaves scope for residual confounding. Variation in disease stage, treatment, sampling time, CALLY thresholds, and model specification further limits comparability. The RFS/PFS synthesis combined clinically distinct endpoints and should not be interpreted as a recurrence-specific estimate. A small number of contributing cohorts also limits the reliability of asymmetry tests and moderator analyses. Associations derived from study-level data do not establish patient-level effect modification, incremental predictive performance, or benefit from CALLY-guided management.

Future work should therefore prioritize prospective, multicenter validation with standardized CALLY calculation, sampling time, cutoffs, endpoint definitions, and covariate adjustment. Serial assessment may be particularly informative. In LA-NSCLC, post-treatment CALLY carried independent prognostic information after thoracic RT, while treatment-related CALLY trajectories were associated with pathological response during neoadjuvant treatment in breast cancer (17, 40). These observations support studying CALLY trajectories rather than relying solely on a single baseline measurement. Future studies should determine whether longitudinal changes improve prediction of treatment response, recovery, recurrence, or survival and whether integration with staging, molecular markers, imaging, or body-composition measures provides clinically meaningful incremental discrimination. Accordingly, our pooled estimates should be interpreted as evidence of prognostic association rather than a validated clinical decision rule.

## Conclusion

Lower CALLY values were associated with poorer survival in the current synthesis of primary lung malignancies. However, the predominantly observational evidence, residual confounding, and variation in CALLY assessment and survival endpoints limit the strength and generalizability of this association. CALLY warrants further evaluation as a readily available prognostic marker, but its incremental value beyond established clinical factors and its ability to improve clinical decisions remain unproven. Prospective multicenter studies should standardize biomarker calculation, sampling time, and outcome definitions and assess external validity before routine clinical implementation.

## Data Availability

All data produced in the present study are available upon reasonable request to the authors
All data produced in the present work are contained in the manuscript

## Abbreviations

AUC: area under the receiver operating characteristic curve
CALLY: C-reactive protein–albumin–lymphocyte
C-index: concordance index
CAR: C-reactive protein-to-albumin ratio
CI: confidence interval
CRC: colorectal cancer
CRP: C-reactive protein
DLBCL: diffuse large B-cell lymphoma
DMFS: distant metastasis-free survival
GRADE: Grading of Recommendations Assessment, Development and Evaluation
HR: hazard ratio
ICI: immune checkpoint inhibitor
IQR: interquartile range
KM: Kaplan–Meier
LA-NSCLC: locally advanced non-small cell lung cancer
LC: lung cancer
LIPI: lung immune prognostic index
LPFS: locoregional progression-free survival
mGPS: modified Glasgow Prognostic Score
MLR: monocyte-to-lymphocyte ratio
MV: multivariable
NLR: neutrophil-to-lymphocyte ratio
NR: not reported
NSCLC: non-small cell lung cancer
OS: overall survival
PFS: progression-free survival
PLR: platelet-to-lymphocyte ratio
PNI: prognostic nutritional index
PRISMA: Preferred Reporting Items for Systematic Reviews and Meta-Analyses
PROSPERO: International Prospective Register of Systematic Reviews
PSM: propensity score matching
QUIPS: Quality In Prognosis Studies
REML: restricted maximum likelihood
RFS: recurrence-free survival
ROC: receiver operating characteristic
RT: radiotherapy
SCLC: small cell lung cancer
SD: standard deviation
SE: standard error
TNM: tumor–node–metastasis
UV: univariable.

## Declarations

### Ethics approval and consent to participate

This systematic review and meta-analysis used aggregate data from published studies and did not involve direct recruitment of participants or access to identifiable individual-level data. Ethics approval and consent to participate were therefore not required.

### Protocol Registration

This study’s protocol has been prospectively registered in PROSPERO (CRD420261475710).

### Consent for publication

Not applicable.

### Availability of data and materials

The data supporting the findings of this systematic review and meta-analysis are available from the corresponding author upon reasonable request.

### Authors’ contributions

Conceptualization: MS, FNR, and DY

Methodology: MS, FNR, PKM, and DY

Software: DY and FNR

Validation: FNR, MS, MY, and AY

Formal Analysis: DY and FNR

Investigation: FNR, MS, DY, IP, and AY

Data Curation: FNR, DY, NN, MY, AY, and MS

Writing – Original Draft: DY, FNR, IP, MY, AR, AY, PKM, NN, and MS

Writing – Review & Editing: MS, DY, FNR, MY, AY, and PKM

Visualization: DY and FNR

Supervision: MS and DY

Project Administration: DY and MS

## Acknowledgements

Not applicable.

